# Assessing clinical acuity in the Emergency Department using the GPT-3.5 Artificial Intelligence Model

**DOI:** 10.1101/2023.08.09.23293795

**Authors:** Christopher Y.K. Williams, Travis Zack, Brenda Y. Miao, Madhumita Sushil, Michelle Wang, Atul J. Butte

## Abstract

This paper evaluates the performance of the Chat Generative Pre-trained Transformer (ChatGPT; GPT-3.5) in accurately identifying higher acuity patients in a real-world clinical context. Using a dataset of 10,000 pairs of patient Emergency Department (ED) visits with varying acuity levels, we demonstrate that GPT-3.5 can successfully determine the patient with higher acuity based on clinical history sections extracted from ED physician notes. The model achieves an accuracy of 84% and an F1 score of 0.83, with improved performance for more disparate acuity scores. Among the 500 pair subsample that was also manually classified by a resident physician, GPT-3.5 achieved similar performance (Accuracy = 0.84; F1 score = 0.85) compared to the physician (Accuracy = 0.86, F1 score = 0.87). Our results suggest that, in real-world settings, GPT-3.5 can perform comparably to physicians on the clinical reasoning task of ED acuity determination.

## Introduction

The November 2022 launch of the Chat Generative Pre-trained Transformer (ChatGPT; GPT-3.5), a general-purpose, 175 billion parameter large language model, has generated widespread attention among researchers, the media and the general public.^1^ Recent studies have already suggested high performance on various natural language tasks, including writing scientific abstracts and achieving a passing score in the United States Medical Licensing Examination.^2,3^ However, these studies are conducted on artificial clinical scenarios, while its performance on real-world clinical text has not been previously evaluated. Determination of clinical acuity, a measure of a patient’s illness severity and the level of medical attention required, is one of the foundational elements of medical reasoning in emergency medicine.^4^ Here, we assess the ability of GPT-3.5 to correctly identify the higher acuity patient, as defined by Emergency Severity Index (ESI), across 10,000 pairs of patients presenting to the Emergency Department.

## Methods

We identified all adult visits to the University of California San Francisco (UCSF) Emergency Department (ED) from 2012 to 2023 with a documented ESI acuity level (range [highest to lowest acuity]: Immediate, Emergent, Urgent, Less Urgent, Non-Urgent) and corresponding ED Physician notes created during the encounter, deidentified and certified as previously described.^5^ From this corpus of deidentified clinical text, regular expressions were used to extract the ‘Chief Complaint’, ‘History of Presenting Illness’ and ‘Review of Systems’ sections from each note which make up a patient’s *Clinical History* (Supplementary File 1). We randomly selected, with replacement, a sample of 10,000 pairs of ED visits with non-equivalent ESI score, balanced for each of the 10 possible pairs of five ESI scores. Using its secure, HIPAA-compliant Application Programming Interface through Microsoft Azure, we queried GPT-3.5 (*gpt-3.5-turbo*) to consider each pair of ED presentations and return which patient was of a higher acuity. A balanced 500 pair subsample was manually classified by a resident physician for comparison of the performance between GPT-3.5 and human classification. The UCSF Institutional Review Board determined that this use of deidentified structured and clinical text data in the UCSF Information Commons is considered non-human-participants research and was exempt from further approval.

## Results

From a total of 251,401 adult Emergency Department visits, we created a balanced sample of 10,000 patient pairs, where each pair contained patients with disparate ESI acuity scores (Supplementary Figure 1). Using only the information documented in the clinical history sections of patients’ first ED physician note, we queried GPT-3.5 to identify the patient with the highest acuity in each pair. Across this sample of paired patient histories, GPT-3.5 correctly inferred the higher acuity patient for 8,354/10,000 pairs (Accuracy = 0.84, F1 score = 0.83). As expected, model performance improved as acuity scores became more disparate between pairs (Table 1), with up to 98% accuracy when distinguishing patients with ‘Immediate’ compared to ‘Less Urgent’ or ‘Non-Urgent’ acuity levels. Among the 500 pair subsample that was also manually classified, GPT-3.5 achieved similar performance (Accuracy = 0.84; F1 score = 0.85) compared to the resident physician (Accuracy = 0.86, F1 score = 0.87) (Figure 1), again using only the clinical history sections of the ED physician note.

**Table 1.** Evaluation of GPT-3.5 performance for each type of Emergency Severity Index (ESI) acuity level pairing: a) Accuracy and b) F1 score.

|  | **Emergency Severity Index (ESI) acuity level** |  |  |  |  |
| --- | --- | --- | --- | --- | --- |
|  | *Immediate* | *Emergent* | *Urgent* | *Less Urgent* | *Non-Urgent* |
| **a) Accuracy** |  |  |  |  |  |
| *Immediate* |  |  |  |  |  |
| *Emergent* | 0.83 |  |  |  |  |
| *Urgent* | 0.93 | 0.71 |  |  |  |
| *Less Urgent* | 0.98 | 0.88 | 0.74 |  |  |
| *Non-Urgent* | 0.98 | 0.92 | 0.81 | 0.58 |  |
| **b) F1 score** |  |  |  |  |  |
| *Immediate* |  |  |  |  |  |
| *Emergent* | 0.83 |  |  |  |  |
| *Urgent* | 0.93 | 0.71 |  |  |  |
| *Less Urgent* | 0.98 | 0.87 | 0.71 |  |  |
| *Non-Urgent* | 0.98 | 0.91 | 0.81 | 0.51 |  |

**Figure 1.**
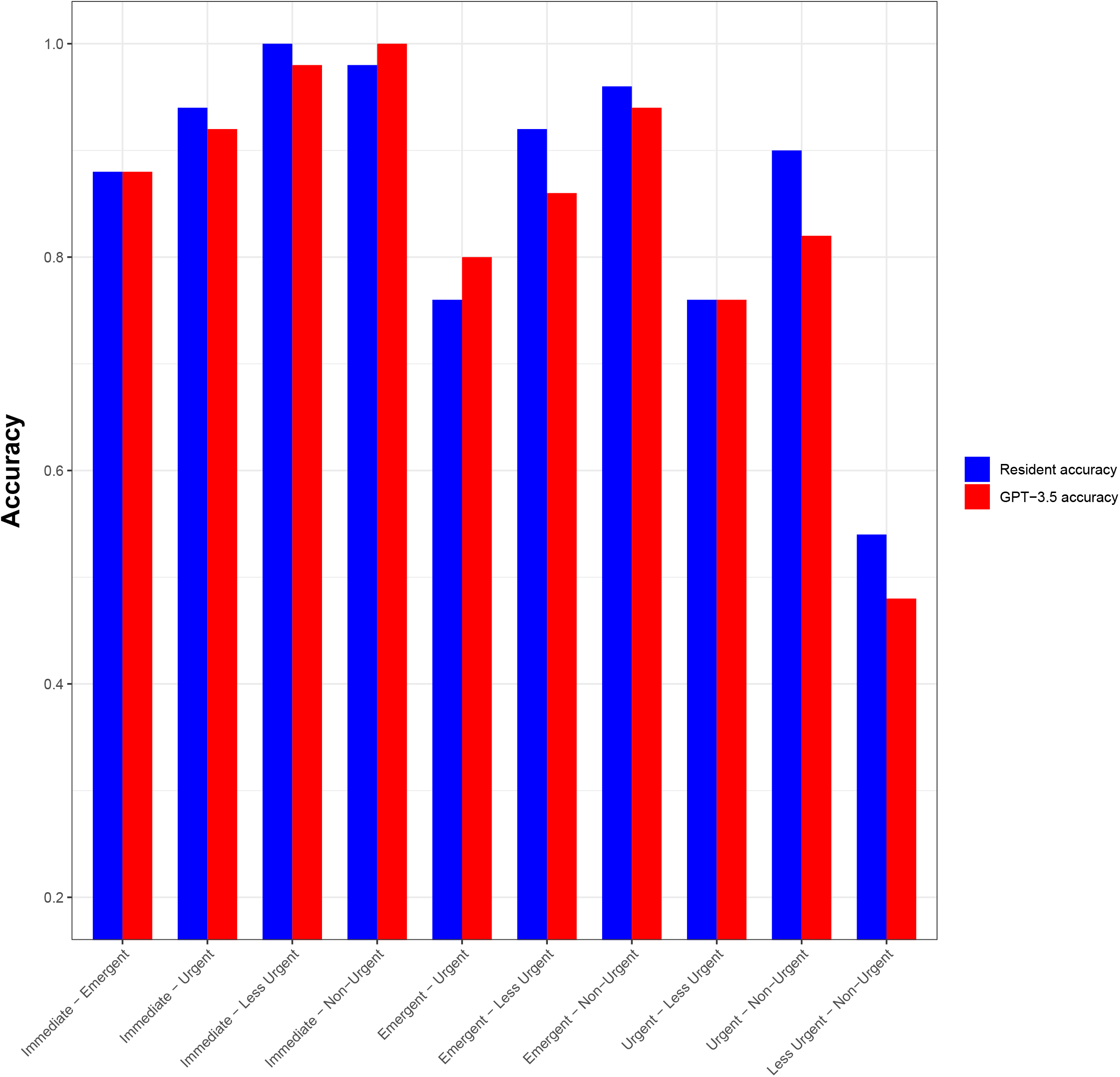
Evaluation of GPT-3.5 (*red*) and resident physician (*blue*) performance for each type of Emergency Severity Index (ESI) acuity level pairing in the 500 pair subsample

## Discussion

This study represents an early and highly powered evaluation of GPT-3.5’s ability to assess real-world clinical text and stratify patients based on their clinical acuity. We found that GPT-3.5 could accurately identify the higher acuity patient when given pairs of presenting histories extracted from patients’ first ED documentation. Among the subsample of patient pairs assessed by both GPT-3.5 and physician, overall performance was comparable. Limitations include the lack of additional prompt engineering to further optimize GPT-3.5 performance, the possibility that ESI scores do not fully represent a patient’s acuity, and the absence of complete details on GPT-3.5 training.^6^ Despite differences in structure and vocabulary between clinical text and more general corpora, our results suggest that, in real-world settings, GPT-3.5 can perform comparably to physicians on the clinical reasoning task of ED acuity determination.

## Data Availability

Data available: No

## Conflicts of Interest

AJB is a co-founder and consultant to Personalis and NuMedii; consultant to Mango Tree Corporation, and in the recent past, Samsung, 10x Genomics, Helix, Pathway Genomics, and Verinata (Illumina); has served on paid advisory panels or boards for Geisinger Health, Regenstrief Institute, Gerson Lehman Group, AlphaSights, Covance, Novartis, Genentech, and Merck, and Roche; is a shareholder in Personalis and NuMedii; is a minor shareholder in Apple, Meta (Facebook), Alphabet (Google), Microsoft, Amazon, Snap, 10x Genomics, Illumina, Regeneron, Sanofi, Pfizer, Royalty Pharma, Moderna, Sutro, Doximity, BioNtech, Invitae, Pacific Biosciences, Editas Medicine, Nuna Health, Assay Depot, and Vet24seven, and several other non-health related companies and mutual funds; and has received honoraria and travel reimbursement for invited talks from Johnson and Johnson, Roche, Genentech, Pfizer, Merck, Lilly, Takeda, Varian, Mars, Siemens, Optum, Abbott, Celgene, AstraZeneca, AbbVie, Westat, and many academic institutions, medical or disease specific foundations and associations, and health systems. AJB receives royalty payments through Stanford University, for several patents and other disclosures licensed to NuMedii and Personalis. AJB’s research has been funded by NIH, Peraton (as the prime on an NIH contract), Genentech, Johnson and Johnson, FDA, Robert Wood Johnson Foundation, Leon Lowenstein Foundation, Intervalien Foundation, Priscilla Chan and Mark Zuckerberg, the Barbara and Gerson Bakar Foundation, and in the recent past, the March of Dimes, Juvenile Diabetes Research Foundation, California Governor’s Office of Planning and Research, California Institute for Regenerative Medicine, L’Oreal, and Progenity. None of these entities had any bearing on the design of this study or the writing of the manuscript.

## Acknowledgements

The authors acknowledge the use of the UCSF Information Commons computational research platform, developed and supported by UCSF Bakar Computational Health Sciences Institute.

## Supplementary Methods

### Cohort selection

Only adult patient (≥18 years) Emergency Department (ED) visits were considered in this study. ED visits with no associated clinical notes were excluded, as were visits with clinical notes written only by non-Emergency Medicine providers. If more than one Emergency Medicine provider note was available for a particular ED visit, the earliest note was selected. In the case of multiple notes with the same chart time, the longest note (by word count) was selected.

### Emergency Severity Index triage system

The Emergency Severity Index (ESI) is the triage system recommended by the American College of Emergency Physicians and Emergency Nurses Association.^1^ It is recorded during the initial triage of patients on presentation to the Emergency Department and provides an indication of how acutely unwell a patient is, how urgently they require medical attention, and the number of anticipated resources required during their encounter. There are 5 acuity levels based on how urgently patients need to be seen by the physician or healthcare provider: immediate, emergent, urgent, less urgent, and non-urgent.^1^ In this study, the ESI was used as the ground-truth indication of which patient presented with a higher clinical acuity, allowing a comparison between GPT-3.5 and human (resident physician) inference.

### Note pre-processing & segmentation

Clinical notes were minimally preprocessed - only new lines and extra spaces were removed. A series of Regular Expressions were used to examine the structure of notes, confirming the presence/absence of the following note headers: ‘Chief Complaint’ (261,688/264,912 notes); ‘Review of Systems’ (261,554/264,912 notes); ‘Physical Exam’ (263,702/264,912 notes); ‘ED Course’ (232,778/264,912 notes); and ‘Initial Assessment’ (186,620/264,912 notes). For each clinical note, we extracted all text from:

1. Clinical History: section ‘Chief Complaint’ (inclusive) to ‘Physical Exam’, representing the full history of each patient’s ED visit, including both their Presenting Complaint/History of Presenting Complaint and Systems Review;
2. Examination: section ‘Physical Exam’ (inclusive) to either ‘ED course’ or ‘Initial Assessment’, representing the Physical Examination findings; and
3. Assessment/Plan: from ‘ED course’ or ‘Initial Assessment’ to note end, representing the clinician’s Impression/Assessment and Plan.

### Tokenisation

A sample of the segmented note text was examined to confirm proper extraction. The dataset was subsequently filtered to remove ED visits with an unspecified ESI acuity score. Only ED visits in which all three sections of the accompanying Emergency Medicine Provider note could be segmented and extracted were included. For this study, only text from the Clinical History section of patients’ clinical notes was analysed by GPT-3.5.

The number of tokens for each section was calculated using the *tiktoken* tokenizer module recommended by Open AI. Tokens can be thought of as pieces of words which form the input of large language models; 100 tokens are approximately equal to 75 words.^2^ Notably, GPT-3.5 has a maximum limit of 4096 tokens shared between prompt (input) and completion (output). Because our prompt required a comparison of Clinical Histories between two different patients presenting to the ED, we further filtered our dataset to remove the minority of ED visits with a Clinical History of greater than 2000 tokens in length.

### Sample selection

Following the creation of this master dataset, we selected, with replacement, a 10,000 pair sample on which GPT-3.5 performance was evaluated. This sample was balanced for each of the 10 paired classes of ESI acuity score:

- 1000 ‘Immediate’ : ‘Emergent’ pairs of ED visits
- 1000 ‘Immediate’ : ‘Urgent’ pairs of ED visits
- 1000 ‘Immediate’ : ‘Less Urgent’ pairs of ED visits
- 1000 ‘Immediate’ : ‘Non-Urgent’ pairs of ED visits
- 1000 ‘Emergent’ : ‘Urgent’ pairs of ED visits
- 1000 ‘Emergent’ : ‘Less Urgent’ pairs of ED visits
- 1000 ‘Emergent’ : ‘Non-Urgent’ pairs of ED visits
- 1000 ‘Urgent’ : ‘Less Urgent’ pairs of ED visits
- 1000 ‘Urgent’ : ‘Non-Urgent’ pairs of ED visits
- 1000 ‘Less Urgent’ : ‘Non-Urgent’ pairs of ED visits

### GPT-3.5 prompt

We used GPT-3.5 to perform zero shot classification of which patient was of a higher acuity based on their Clinical History. Using Regular Expressions, we confirmed that there was no mention of a patient’s acuity level in their Clinical History to ensure no data leakage would confound our results. We deployed the following template for prompting GPT-3.5, with Patient A and Patient B representing the two Clinical Histories for any particular pair of ED visits:

*You are an Emergency Department physician. Below are the symptoms of two different patients presenting to the Emergency Department, Patient A and Patient B. Please return which patient is of the highest acuity between these two patients. Please return one of two answers: ‘0: Patient A is of higher acuity’ ‘1: Patient B is of higher acuity’ Please do not return any additional explanation*.

*Patient A: “ “*

*Patient B: “ “*

This template was chosen following several rounds of prompt engineering to ensure that only the two stated outputs (‘0: Patient A is of higher acuity’ or ‘1: Patient B is of higher acuity’) were returned by the model. This was necessary as GPT-3.5 has a tendency to return verbose answers which otherwise would be difficult to analyse at scale. We did not conduct additional prompt engineering to further improve model performance.

We randomly shuffled whether patient A or B was the higher acuity patient to prevent possible systemic bias in the way GPT-3.5 returns a response from confounding our results (e.g if GPT-3.5 is more likely to return ‘Patient A’ as its response, regardless of the Clinical History given).

### GPT-3.5 and human evaluation

Prompts were sent to the GPT-3.5 Application Programming Interface (API) (model = ‘gpt-3.5-turbo-0301’, role = ‘user’, temperature = 0; all other settings at default values) via the HIPAA-compliant, UCSF Secure Azure OpenAI environment and responses from the API were recorded. The higher acuity patient (‘A’ or ‘B’) was extracted from the API output using Regular Expressions and compared to the ground-truth acuity level. Separately, a resident physician blinded to both the GPT-3.5 labels and ground-truth labels reviewed the Clinical Histories of a balanced 500 pair subsample (n = 50 for each of the 10 categories) to determine which of Patient A or B was of the higher acuity. Accuracy and binary F1 scores were calculated for both GPT-3.5 and human annotator for comparison.

**Supplementary Figure 1.**
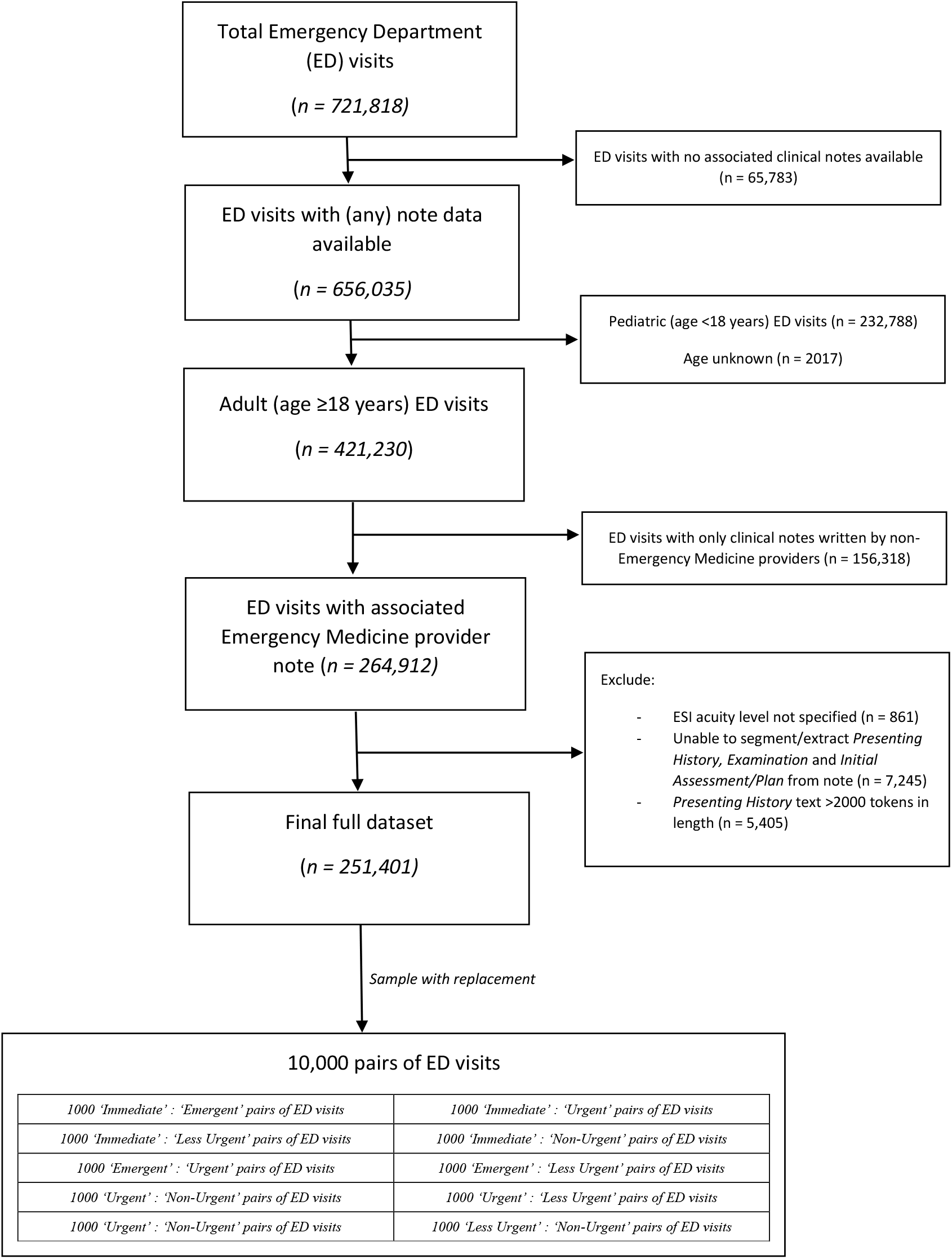
Flowchart of included Emergency Department visits and construction of 10,000 pair sample

## Notes

### Funding Statement

This study did not receive any funding.

### Author Declarations

The UCSF Institutional Review Board determined that this use of deidentified structured and clinical text data in the UCSF Information Commons is considered non-human-participants research and was exempt from further approval.

